# News from the front: Estimation of excess mortality and life expectancy in the major epicenters of the COVID-19 pandemic in Italy

**DOI:** 10.1101/2020.04.29.20084335

**Authors:** Simone Ghislandi, Raya Muttarak, Markus Sauerberg, Benedetta Scotti

**Author notes:** **Corresponding authors:** Raya Muttarak, Address: International Institute for Applied Systems Analysis, Schlossplatz 1, A-2361 Laxenburg, Austria. **Author Contributions** S.G. and B.S. designed research; B.S. collected and compile data; B.S. and M.S. performed research and analysed data; S.G., R.M., M.S. and R.M. interpreted the results and wrote the paper.

## Abstract

Existing studies on the mortality impacts of the COVID-19 pandemic commonly rely on national official reports. However, in a pandemic, deaths from COVID-19 can be miscounted due to under-reporting and inaccurate death registration. Official statistics on COVID-19 mortality are sensitive to classification, estimation and reporting practice which are not consistent across countries. Likewise, the reported mortality is often provided at the national level which results in underestimation of the true scale of the human life impact given that the outbreaks are localised.

This study overcomes the problem of under-reporting of COVID-19-related deaths by using all cause daily death registrations data provided by the Italian Statistical Office (ISTAT) from January 1 to April 30, 2020 in comparison with official figures reported by the Civil Protection Department. The study focuses on the five most severely hit provinces in Italy (Bergamo, Brescia, Cremona, Lodi and Piacenza) and Lombardy region. We calculate excess mortality in 2020 compared to the average of the years 2015 to 2019 and estimate life expectancy for the first wave of the epidemic and for the rest of the year 2020. Not only is life expectancy a reliable measure of a country’s health status and development, it also allows us to quantify the impact of COVID-19 on human life.

The estimated excess deaths show significantly higher mortality than COVID-19 official mortality statistics, particularly during the peak of the epidemic and amongst people aged 60 years and over. We find that for the first wave of the epidemic, life expectancy in the five provinces reduced by 5.1 to 7.8 and 3.2 to 5.8 years for men and women, respectively. For annual life expectancy for the year 2020, in a scenario with no harvesting effect i.e. mortality rates resume to an average level of the years 2015-2019 after the end of the first epidemic wave, the years of life lost is equivalent to 2 to 3.5 years for men and 1.1. to 2.5 years for women in the five provinces.

The COVID-19 pandemic posed a substantial impact on population health in Italy as it represents the largest decline in life expectancy after the 1918 influenza pandemic and the Second World War.

## Introduction

Italy is the first country in Europe to experience a large scale impact of COVID-19 pandemic on human life. As the COVID-19 outbreak spread aggressively to the rest of the world, disparities in COVID-19 deaths became distinct. Comparing COVID-19 deaths across countries, however, is challenging. Most of the existing literature has relied on case-fatality rates (CFR) as a measure of mortality ^1–4^. However, CFR are not informative for international and historical comparisons. Since they are calculated as the number of deaths divided by the number of confirmed cases, the absence of accurate estimation of the infection rates in a reference population makes the denominator in the CFR reliant on testing strategies and capacities ^5^.

In addition, there is not a uniform way of classifying, recording and reporting COVID-19 deaths^6^. Italy, for instance, classifies as an official COVID 19 decease everyone who tested positive for the virus before death, regardless of the pre-existing diseases ^4^. Likewise, while countries like Belgium and France (after 2 April) include coronavirus deaths outside hospitals in their official daily reports, countries like the UK, Italy or Spain do not. Some countries such as Belgium, the UK and the US record COVID-19 deaths also from suspected cases and do not require a positive laboratory diagnosis unlike in Germany and the Netherlands ^7^. In addition, when the epidemic worsens, the counting of fatalities becomes more difficult. People dying at home or in long-term care facilities might indeed not be tested at all simply because resource allocation prioritizes emergency operations ^8–10^. The overall number of COVID-19 deaths thus is most likely undercounted.

The COVID-19 pandemic can also have an indirect impact on human life. As medical resources are primarily allocated for the treatment of COVID-19 patients, non-COVID medical emergencies and cares for chronic illness are likely to be undermined ^11^. Depleted hospital capacity in providing routine medical treatments, coupled with reluctance to visit healthcare facilities due to a fear of the spread of COVID-19 infection within the hospital, could contribute to peaks in mortality for other medical conditions that are not directly related to the epidemic ^12^. Recent evidence shows that the incidence of out-of-hospital cardiac arrest, for instance, has increased in Italy ^13^ and France ^14^ as compared to the same period in the previous years. Arguably, these deaths should also be considered as death indirectly related to COVID-19.

To overcome the problems related to both the variation in mortality measurements and the indirect effects of the pandemic, this study uses daily death registrations data for the period 1 January to 30 April 2020 published by the Italian Statistical Office (ISTAT). This allows us to calculate excess all-cause mortality during the period of the epidemic and estimate the corresponding impact on life expectancy without suffering from selection bias related to sampling in testing and reporting policies.

Another contribution of this study is in its specific focus on small-area estimation of mortality and life expectancy. In modelling the spread of COVID-19 in Italy, Gatto et al.^15^ highlight the importance of considering a spatial nature of the epidemic wave. The highly clustered nature of local transmission results in high concentrations of severe illnesses and deaths in one area ^16^. Hence, the direct impact of COVID-19 on mortality rates and average life expectancy is likely to be felt at a sub-national level rather than in a whole country. For this reason, the present study focuses on specific geographical areas in Italy: four provinces in Lombardy (Bergamo, Lodi, Cremona, Brescia) and one province in Emilia Romagna (Piacenza). These represent the hardest hit provinces in the country in terms of excess mortality.

The period of observation of this study is also crucial. Unlike previous studies on excess mortality from COVID-19 ^17,18^, our study includes the entire first wave of COVID-19 epidemic. In particular, the analysis relies exclusively on hard data without any modelling of the future evolution of the mortality wave. As shown in Figure 2, by 30 April 2020 the excess mortality had dropped to zero in all the studied provinces in Italy marking the completion of the first epidemic wave.

Based on daily death registrations data, the present study: i) compares mortality rates in 2015-2019 and 2020 *across age and sex categories*; ii) provides estimations of the changes in life expectancy following the COVID-19 pandemic. This study focuses on the concept of *excess mortality*, defined as the increase in the age-sex-province specific number of deaths in 2020 with respect to the average of the previous five years. These specific figures provide an accurate picture of the impact of the epidemic when compared to “normal” times, showing the differential effects across age groups, sexes and geographical areas. In order to assess the overall cost in terms of human life, however, one needs to further collapse the increases of mortality rates into an index universal enough to provide a reliable measure of human life (years) lost. In this regard, life expectancy is the natural candidate for such an assessment, since it is significantly related to the overall wellbeing of populations and could thus provide a simple, objective and immediate measure of the human casualties associated to unprecedented shocks such as the COVID-19 pandemic ^19–21^. Furthermore, reliable measures of life expectancy are available since the nineteenth century for some countries. This makes it possible to use life expectancy for historical comparisons of the human costs associated to major disruptive events.

### Institutional and geographical context of the hardest hit areas

In the early morning of 21 February 2020, the first severe case of local transmission was diagnosed in Europe, in a small hospital in Codogno, a municipality in the province of Lodi, southeast of Milan ^22^. Initially, authorities reacted by tracing the connections of *patient one* but finally failed to identify a *patient zero*. 11 municipalities within the province of Lodi were put under strict measures to contain the spread of the disease and declared the quarantine “Red Zone” as early as 24 February 2020. Meanwhile, another cluster of COVID-19 outbreak emerged in Alzano Lombardo and Nembro, two municipalities in the province of Bergamo, north east of Milan. Amidst the rapid rise in the number of detected cases, especially in the municipalities surrounding these two epicenters, on 8 March the Italian government imposed a (partial) nationwide lockdown from 9 March, followed by a total lockdown of all non-essential activities on 23 March 2020 ^23^. Whilst the Italian government was praised by WHO for an implementation of such a drastic measure (not being employed in modern democratic nations since World War II), the virus had already spread undetected in the northern part of the country since late January 2020 ^24–26^. The containment measures thus might have been imposed a little too late ^27^. As a consequence, the outbreak has put unprecedented burden on the Italian healthcare system, resulting in an exceptionally high number of coronavirus deaths, currently the fourth highest in the world after the US, Brazil and the UK ^28^.

Geographically, Lodi and Codogno are close to other two provinces included in our sample, Cremona and Piacenza (see Figures S1. and S2. in SI Appendix for a geographical location of the provinces being studied). The epidemic wave involving these provinces is thus considered as part of the Lodi-Codogno cluster. Bergamo and Brescia are located north-east of Milan and, despite the first COVID-19 severe cases being detected just one day after *patient one* in Lodi, experienced a week delay in the arrival of the epidemic wave ^23^. Lombardy, on the other hand, is the most populated region in Italy and the one with the highest Gross Domestic Product (GDP). Overall, it accounts for one sixth of the Italian population and one fifth of its GDP. Lombardy is relevant for our analysis because it is by far the hardest hit region by the COVID-19 pandemic in Italy, accounting for almost 50% of the human casualties of the entire country^5^. Indeed, with the exception of Piacenza (locating in Emilia Romagna region), a bordering province with Lombardy region, all other four provinces included in the analysis are in Lombardy. In the following, we will thus present statistics also for the region of Lombardy.

## Data and Methods

We rely on a compendium of administrative data provided by the Italian National Institute of Statistics (ISTAT). We use daily death counts for all causes at the municipality level, disaggregated by age and sex. For the calendar year 2020, they cover the period between 1 January 1 and 30 April. For the calendar years 2015-2019, they cover the period between 1 January and 31 December. Importantly, ISTAT has not released data on daily death counts for 2020 for all Italian municipalities. These data are available for 7,270 municipalities, out of a total of 7,904 municipalities in Italy, which have been included in the recently established National Register of Resident Population (ANPR). In our geographical areas of interest, the number of municipalities included in the ANPR ranges from 96.7% in the province of Lodi to 100% in the province of Piacenza. For Lombardy, the ANPR coverage is 97.3%.

In addition, we use three further sources of information. First, we obtain data on resident population at the municipality level, disaggregated by sex and single-year age class on 1 January for the years 2015-2019. We reclassify age classes to five-year age group so as to match the ones used by ISTAT for daily and annual death counts and aggregate data accordingly. Second, we use data on monthly live births, disaggregated by sex, at the municipality level. These data are available from January 2015 up to December 2019. Finally, we collect data on monthly death counts, disaggregated by sex, at the municipality level. These data are available from January 2015 up to December 2019.

Methodologically, the estimation approach is standard. Excess number of deaths at any day in 2020 is measured as the difference between the observed and the average number of deaths in the province for the same day in 2015-2019. Age-sex-province specific mortality rates are then applied and standard period life tables are built. Details of the estimation procedure are provided in the Supplemental Materials.

Life expectancy is calculated for two different reference periods. The first quadrimester life expectancy, i.e. life expectancy for the first four months of the year, and the period (annual) life expectancy, i.e. for the entire calendar year. While the former measure does not require any assumption and relies entirely on observed data, annual life expectancy needs assumptions on mortality trends for the rest of the year after 30 April when the available ISTAT data end.

For this reason, we provide two scenarios for the annual life expectancy. In the ‘optimistic’ scenario, harvesting (i.e. the reduction in mortality rates following a temporary peak in mortality associated to negative shocks) is complete and the overall mortality rates by age at the end of the year are kept constant with respect to the previous five years. This approach implies by construction that everyone who died during COVID-19 would have died anyway during the year. In this setting, the main effect of COVID-19 is thus to anticipate mortality to the first quadrimester of the year 2020. Although this scenario is unlikely, it represents a clearly defined upper bound for life expectancy in 2020.

In the ‘business-as-usual’ scenario, on the contrary, harvesting is set to zero and we assume that after the end of the epidemic, mortality rates will go back to the average levels recorded in the previous five years. There are good reasons to think that mortality will not reduce substantially after the end of the first wave of COVID-19 infections. For example, since most of the resources and health services have been directed to COVID-19 patients, people with other medical conditions such as cancer were under-treated. Likewise, although most vulnerable people have been doing self-isolation and locking themselves inside, it is highly possible that they would get the virus later after restriction measures are lifted. As a matter of fact, premature mortality after the first epidemic wave might actually ***increase*** as a consequence of the lockdowns and the following economic recession, as has been observed, for example, in Greece ^29^ and elsewhere in Europe after the 2008 global economic crisis ^30,31^. Furthermore, in some provinces like Bergamo, the number of deaths of males in some age groups by 30 April has already *exceeded the annual number of age-sex specific deaths* in the same provinces in 2019 (Figures S3A and S3B in SI Appendix). The ‘business-as-usual’ scenario therefore is likely to be more realistic than the ‘optimistic’ scenario.

## Results

The geographical distribution of excess deaths in the observed period across Italy (Figure 1) matches with the distribution of confirmed cases (which comprise the deceased, recovered individuals and active cases) provided by the Italian Civil Protection Department, the official surveillance data on COVID-19 ^32^. This geographical pattern warrants that the excess mortality observed in our data represents mortality directly and indirectly related to COVID-19.

**Figure 1:**
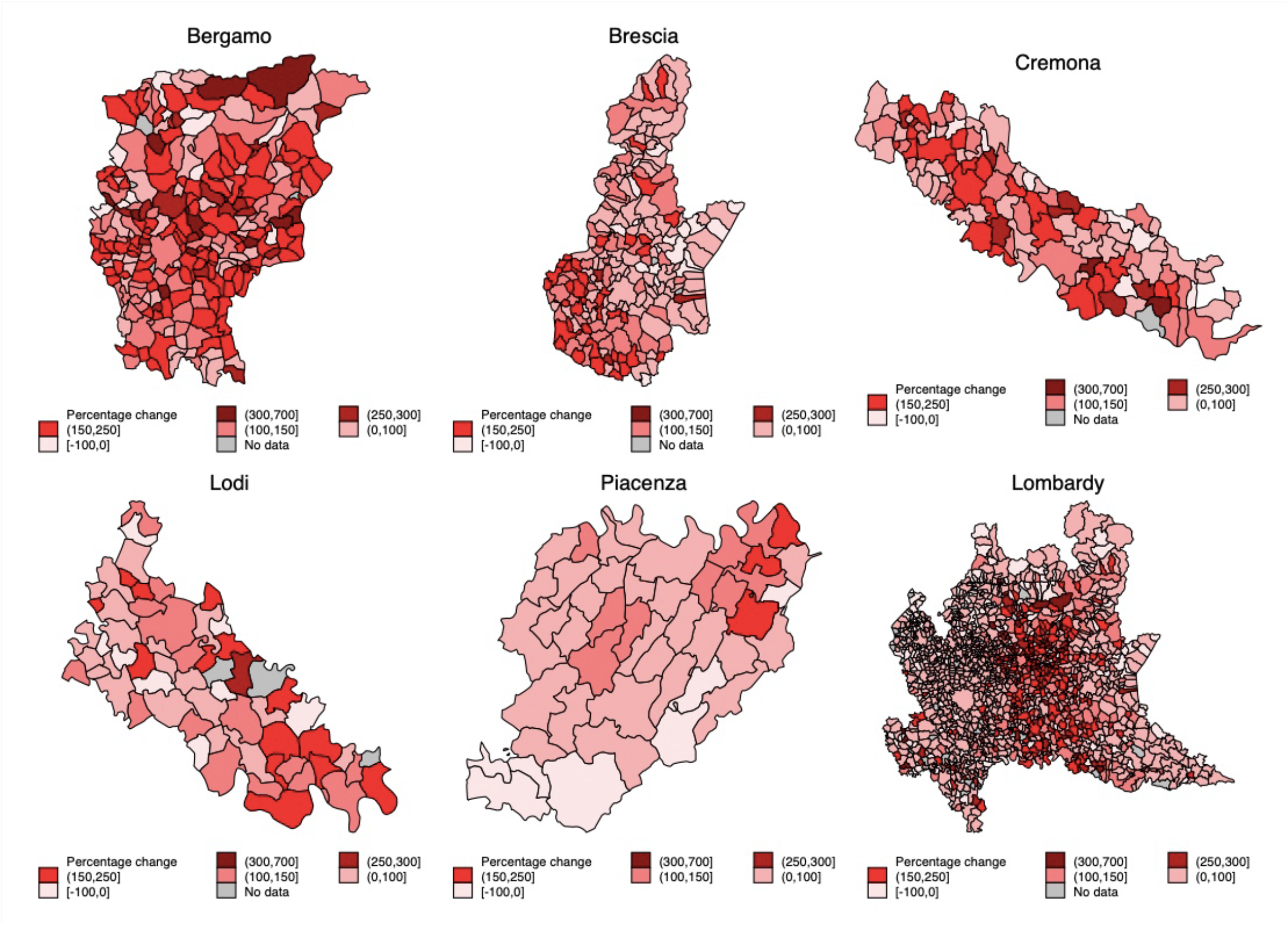
Excess mortality in Italy by municipality in (A) Bergamo, (B) Brescia, (C) Cremona, (D) Lodi, (E) Piacenza and (F) Lombardy. The maps plot the percentages change in total number of deaths recorded between 1 January – 30 April 2020 with respect to the 2015-2019 average.

**Figure 2:**
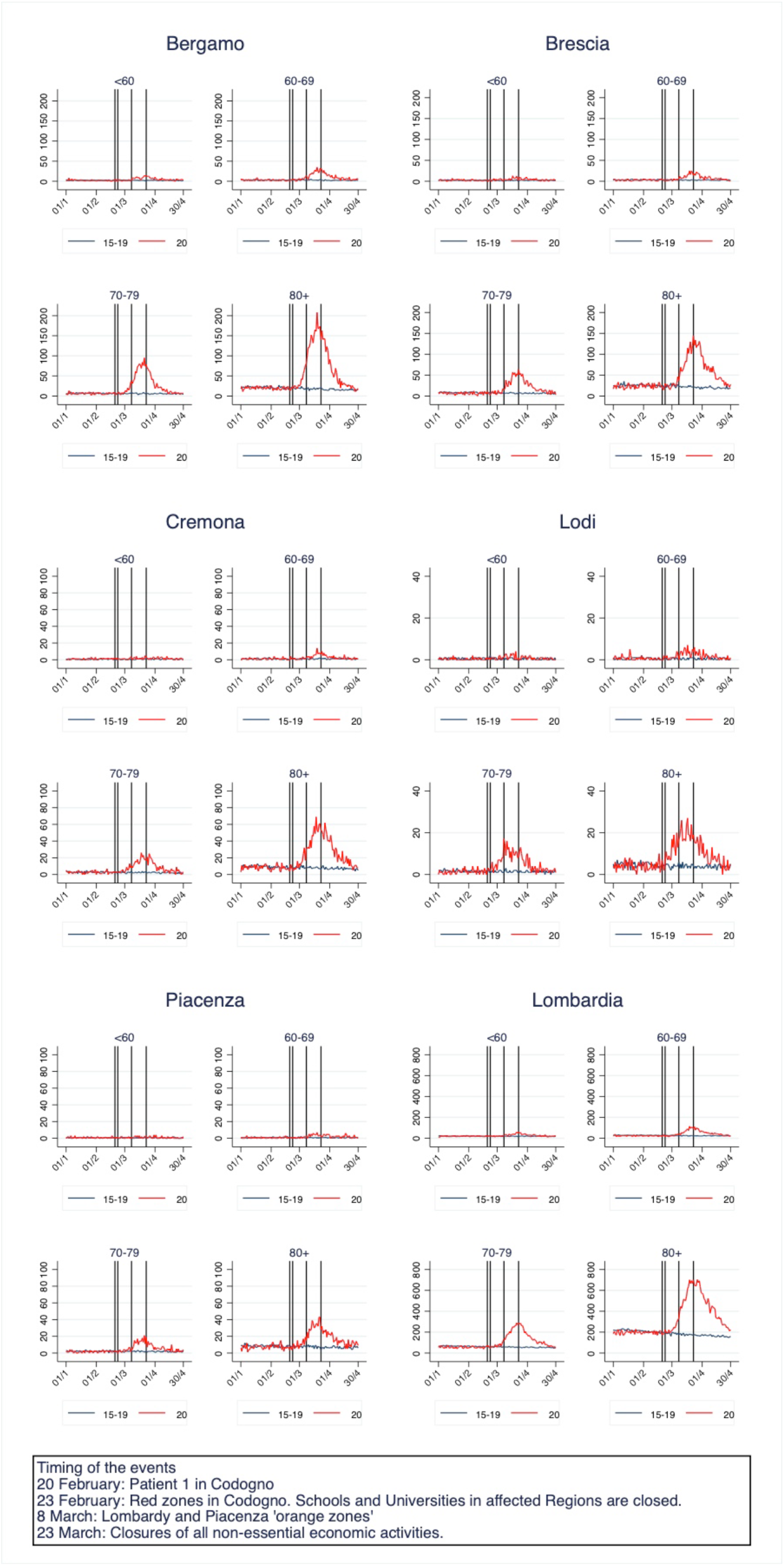
Trends in daily mortality by age in selected five provinces and in Lombardy: (A) Bergamo, (B) Brescia, (C) Cremona, (D) Lodi, (E)Piacenza and (F) Lombardy. The graphs plot the total daily death count between 1 January and 30 April (2020 vs 2015-2019 average), males and females combined. The vertical lines show relevant dates for the evolution of the epidemic. Vertical lines indicate relevant days: 20 February=Patient 1 found in Codogno; 23 February= Red zones in Codogno. Schools and Universities in affected Regions are closed; 8 March = Lombardy and Piacenza orange zones; 23 March = Closures of all non-essential economic activities.

Bergamo, Brescia, Cremona, Lodi and Piacenza are the top five provinces in Italy experiencing the highest increase in mortality in the first quadrimester. Compared to the average number of people that died in the same period in the previous five years (2015-2019), the excess number of deaths (for those aged over 40) between 1 January and 30 April 2020 sums to 5980 in Bergamo, 4196 in Brescia, 1996 in Cremona, 897 in Lodi and 1127 in Piacenza. For the entire region of Lombardy, the excess number of deaths are approximately 23,363 (Table 1). Mortality rate in the first quadrimester of 2020 increased substantially in all provinces for all age groups, with the highest increase being for men aged 70-79 in Bergamo (a 380% increase). Age clearly represents a risk factor for excess mortality in a similar manner as age-gradient in COVID-19 CFR found in Italy and elsewhere ^33^. Among the excess deaths observed in Bergamo, for instance, mortality rate is much higher among older men aged ≥ 70 years (16 times higher than those aged below 70). A similar ratio is found in other provinces.

**Table 1:**
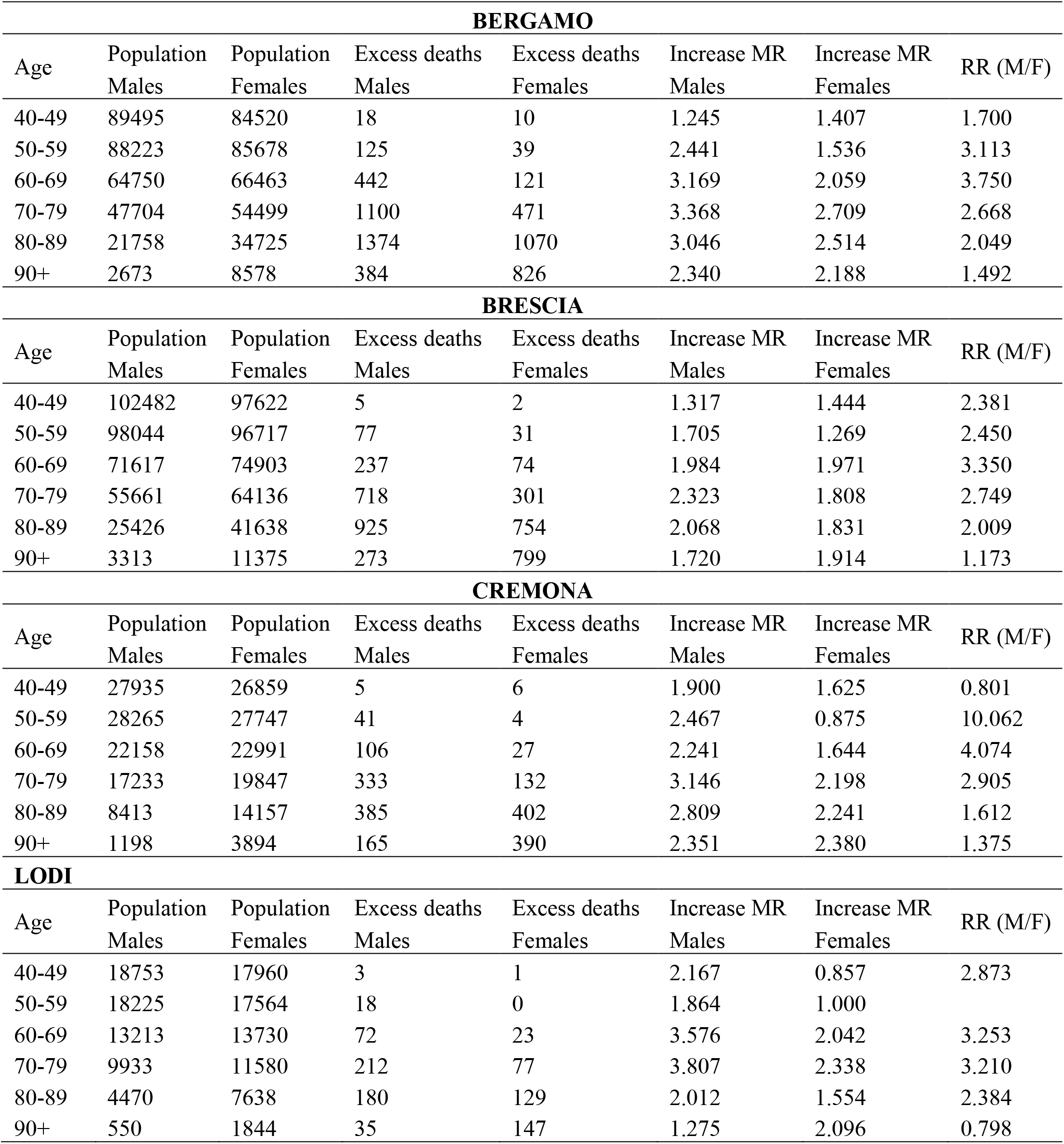
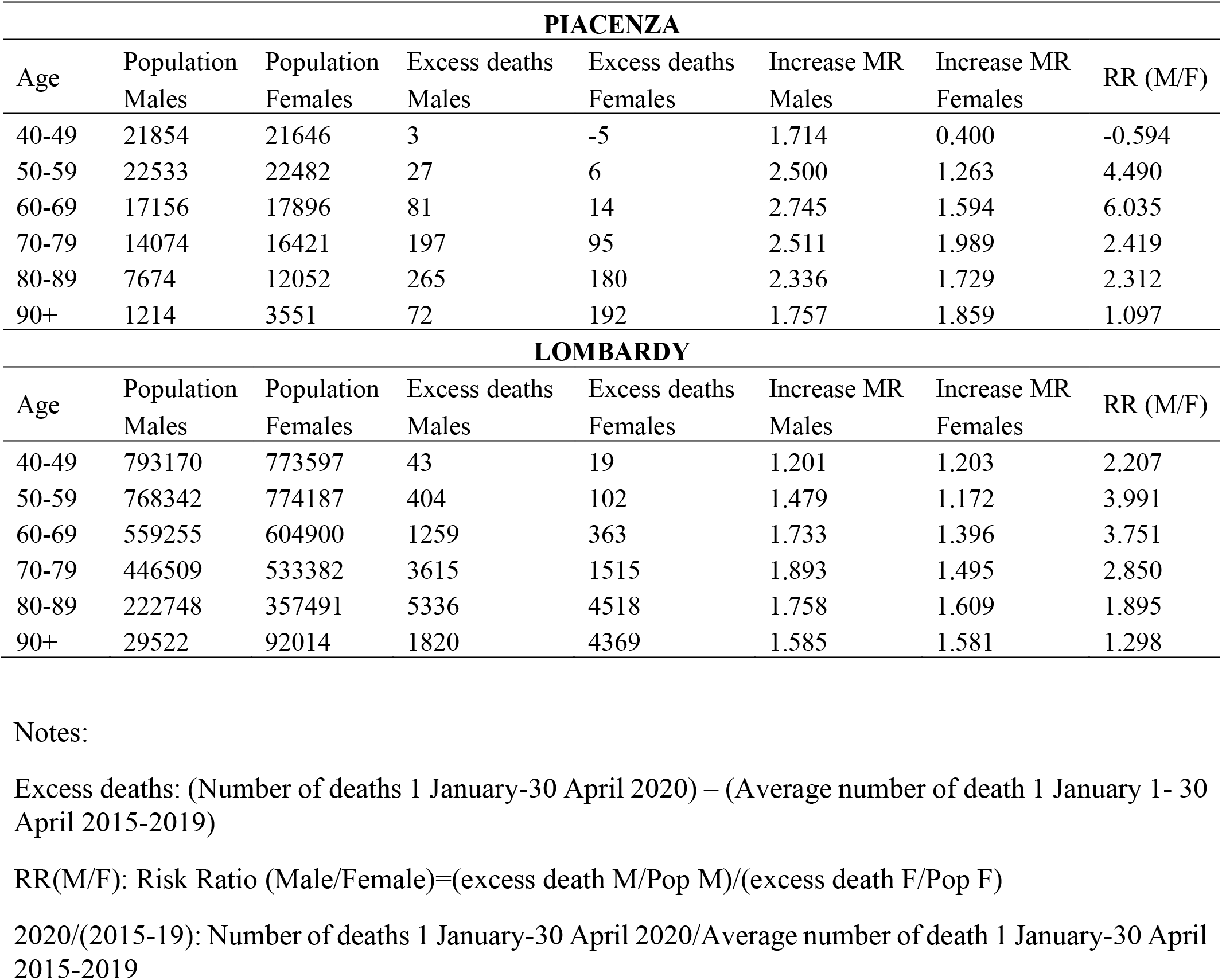
Total population, excess mortality and male to female relative risk (RR) by province, age and sex

| **BERGAMO** |  |  |  |  |  |  |  |
| --- | --- | --- | --- | --- | --- | --- | --- |
| Age | Population<br>Males | Population<br>Females | Excess deaths<br>Males | Excess deaths<br>Females | Increase MR<br>Males | Increase MR<br>Females | RR (M/F) |
| 40-49 | 89495 | 84520 | 18 | 10 | 1.245 | 1.407 | 1.700 |
| 50-59 | 88223 | 85678 | 125 | 39 | 2.441 | 1.536 | 3.113 |
| 60-69 | 64750 | 66463 | 442 | 121 | 3.169 | 2.059 | 3.750 |
| 70-79 | 47704 | 54499 | 1100 | 471 | 3.368 | 2.709 | 2.668 |
| 80-89 | 21758 | 34725 | 1374 | 1070 | 3.046 | 2.514 | 2.049 |
| 90+ | 2673 | 8578 | 384 | 826 | 2.340 | 2.188 | 1.492 |
| **BRESCIA** |  |  |  |  |  |  |  |
| Age | Population<br>Males | Population<br>Females | Excess deaths<br>Males | Excess deaths<br>Females | Increase MR<br>Males | Increase MR<br>Females | RR (M/F) |
| 40-49 | 102482 | 97622 | 5 | 2 | 1.317 | 1.444 | 2.381 |
| 50-59 | 98044 | 96717 | 77 | 31 | 1.705 | 1.269 | 2.450 |
| 60-69 | 71617 | 74903 | 237 | 74 | 1.984 | 1.971 | 3.350 |
| 70-79 | 55661 | 64136 | 718 | 301 | 2.323 | 1.808 | 2.749 |
| 80-89 | 25426 | 41638 | 925 | 754 | 2.068 | 1.831 | 2.009 |
| 90+ | 3313 | 11375 | 273 | 799 | 1.720 | 1.914 | 1.173 |
| **CREMONA** |  |  |  |  |  |  |  |
| Age | Population<br>Males | Population<br>Females | Excess deaths<br>Males | Excess deaths<br>Females | Increase MR<br>Males | Increase MR<br>Females | RR (M/F) |
| 40-49 | 27935 | 26859 | 5 | 6 | 1.900 | 1.625 | 0.801 |
| 50-59 | 28265 | 27747 | 41 | 4 | 2.467 | 0.875 | 10.062 |
| 60-69 | 22158 | 22991 | 106 | 27 | 2.241 | 1.644 | 4.074 |
| 70-79 | 17233 | 19847 | 333 | 132 | 3.146 | 2.198 | 2.905 |
| 80-89 | 8413 | 14157 | 385 | 402 | 2.809 | 2.241 | 1.612 |
| 90+ | 1198 | 3894 | 165 | 390 | 2.351 | 2.380 | 1.375 |
| **LODI** |  |  |  |  |  |  |  |
| Age | Population<br>Males | Population<br>Females | Excess deaths<br>Males | Excess deaths<br>Females | Increase MR<br>Males | Increase MR<br>Females | RR (M/F) |
| 40-49 | 18753 | 17960 | 3 | 1 | 2.167 | 0.857 | 2.873 |
| 50-59 | 18225 | 17564 | 18 | 0 | 1.864 | 1.000 |  |
| 60-69 | 13213 | 13730 | 72 | 23 | 3.576 | 2.042 | 3.253 |
| 70-79 | 9933 | 11580 | 212 | 77 | 3.807 | 2.338 | 3.210 |
| 80-89 | 4470 | 7638 | 180 | 129 | 2.012 | 1.554 | 2.384 |
| 90+ | 550 | 1844 | 35 | 147 | 1.275 | 2.096 | 0.798 |

| **PIACENZA** |  |  |  |  |  |  |  |
| --- | --- | --- | --- | --- | --- | --- | --- |
| Age | Population<br>Males | Population<br>Females | Excess deaths<br>Males | Excess deaths<br>Females | Increase MR<br>Males | Increase MR<br>Females | RR (M/F) |
| 40-49 | 21854 | 21646 | 3 | -5 | 1.714 | 0.400 | -0.594 |
| 50-59 | 22533 | 22482 | 27 | 6 | 2.500 | 1.263 | 4.490 |
| 60-69 | 17156 | 17896 | 81 | 14 | 2.745 | 1.594 | 6.035 |
| 70-79 | 14074 | 16421 | 197 | 95 | 2.511 | 1.989 | 2.419 |
| 80-89 | 7674 | 12052 | 265 | 180 | 2.336 | 1.729 | 2.312 |
| 90+ | 1214 | 3551 | 72 | 192 | 1.757 | 1.859 | 1.097 |
| **LOMBARDY** |  |  |  |  |  |  |  |
| Age | Population<br>Males | Population<br>Females | Excess deaths<br>Males | Excess deaths<br>Females | Increase MR<br>Males | Increase MR<br>Females | RR (M/F) |
| 40-49 | 793170 | 773597 | 43 | 19 | 1.201 | 1.203 | 2.207 |
| 50-59 | 768342 | 774187 | 404 | 102 | 1.479 | 1.172 | 3.991 |
| 60-69 | 559255 | 604900 | 1259 | 363 | 1.733 | 1.396 | 3.751 |
| 70-79 | 446509 | 533382 | 3615 | 1515 | 1.893 | 1.495 | 2.850 |
| 80-89 | 222748 | 357491 | 5336 | 4518 | 1.758 | 1.609 | 1.895 |
| 90+ | 29522 | 92014 | 1820 | 4369 | 1.585 | 1.581 | 1.298 |

Notes:
Excess deaths: (Number of deaths 1 January-30 April 2020) – (Average number of death 1 January 1- 30 April 2015-2019)
RR(M/F): Risk Ratio (Male/Female)=(excess death M/Pop M)/(excess death F/Pop F)
2020/(2015-19): Number of deaths 1 January-30 April 2020/Average number of death 1 January-30 April 2015-2019

When simply considering the distribution of excess mortality without adjusting for population size in each age-sex category, we observe slightly more excess mortality in men than in women (53% of excess deaths involve male subjects). However, when considering the mortality risk ratio between sexes, the excess mortality for males is consistently higher than that of females across all age groups and provinces (relative risk >1). For example, 70-79 years old men in Bergamo were 2.75 times more likely to die than women of the same age. It is evident that male sex is a risk factor associated with severe illness and deaths from COVID-19 in Italy ^34^ and elsewhere ^35,36^. Our evidence on excess mortality suggests similar sex disparities in COVID-19 related deaths.

Figure 2 shows trends in daily mortality for the five provinces and Lombardy. Plotting mortality distribution by age groups allows us to fully capture the progression of the epidemic. It is evident how the epidemic curve inflates with age across all provinces. It is also clear that by 30 April the daily mortality in all selected provinces approached the pre-pandemic values (i.e. with no excess mortality). Hence, the wavelength of the epidemic in these provinces was between 6 and 8 weeks, with the peak happening around two weeks after the onset of the outbreak.

Vertical lines show four relevant dates for the evolution of the epidemic. With the case of *patient one* being first identified in Codogno located in the province of Lodi, the authorities quickly locked down 11 municipalities in the area on 24 February 2020. The containment measures through lockdown were only implemented in other provinces from March 8 on. Although the earlier lockdown enabled Lodi to flatten the curve more effectively than other severely hit provinces (Figure 2), the province still experienced a notable increase in excess mortality. Considering that the incubation period – the time between exposure and symptom onset – can be up to 24 days ^37,38^, it is evident that the lockdown were imposed too late in these provinces. Apart from political reasons preventing the authorities to introduce the lockdown earlier in the provinces where the number of cases had been risen rapidly like Bergamo, the recent evidence shows that COVID-19 has already been circulated undetected in northern Italy since January 2020 ^24–26^. Our study thus proxies the impact of COVID-19 outbreak in the absence of containment interventions.

Trends in first quadrimester and annual life expectancies are illustrated in Figure 3 and Figure 4.

**Figure 3:**
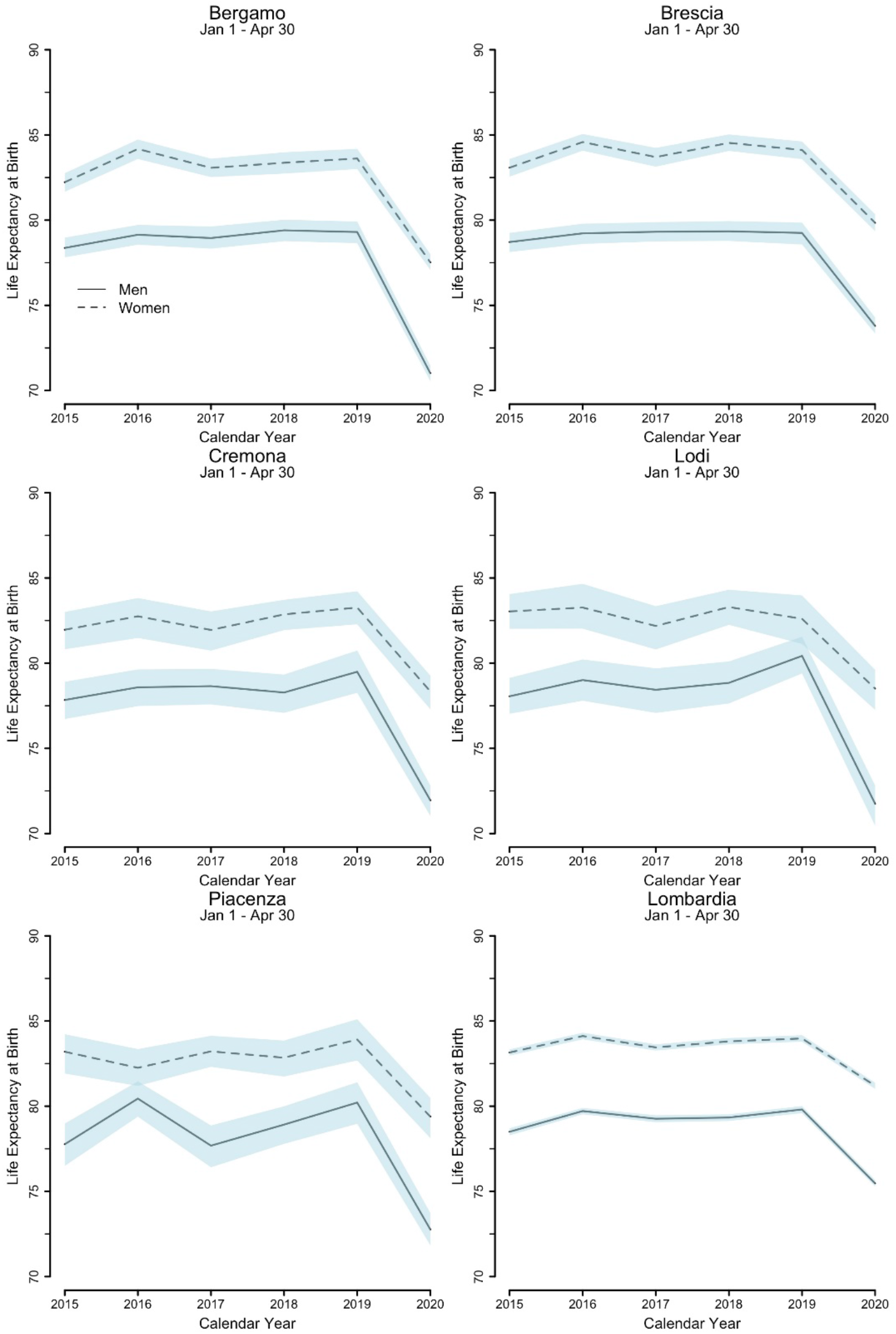
Estimates of the first quadrimester (1 January – 3 April 30) life expectancy at birth by sex in selected provinces and in Lombardy. Confidence intervals (95%) for life expectancies are estimated by bootstrapping using Monte Carlo simulation methods, assuming death counts follow a binomial distribution.

**Figure 4.**
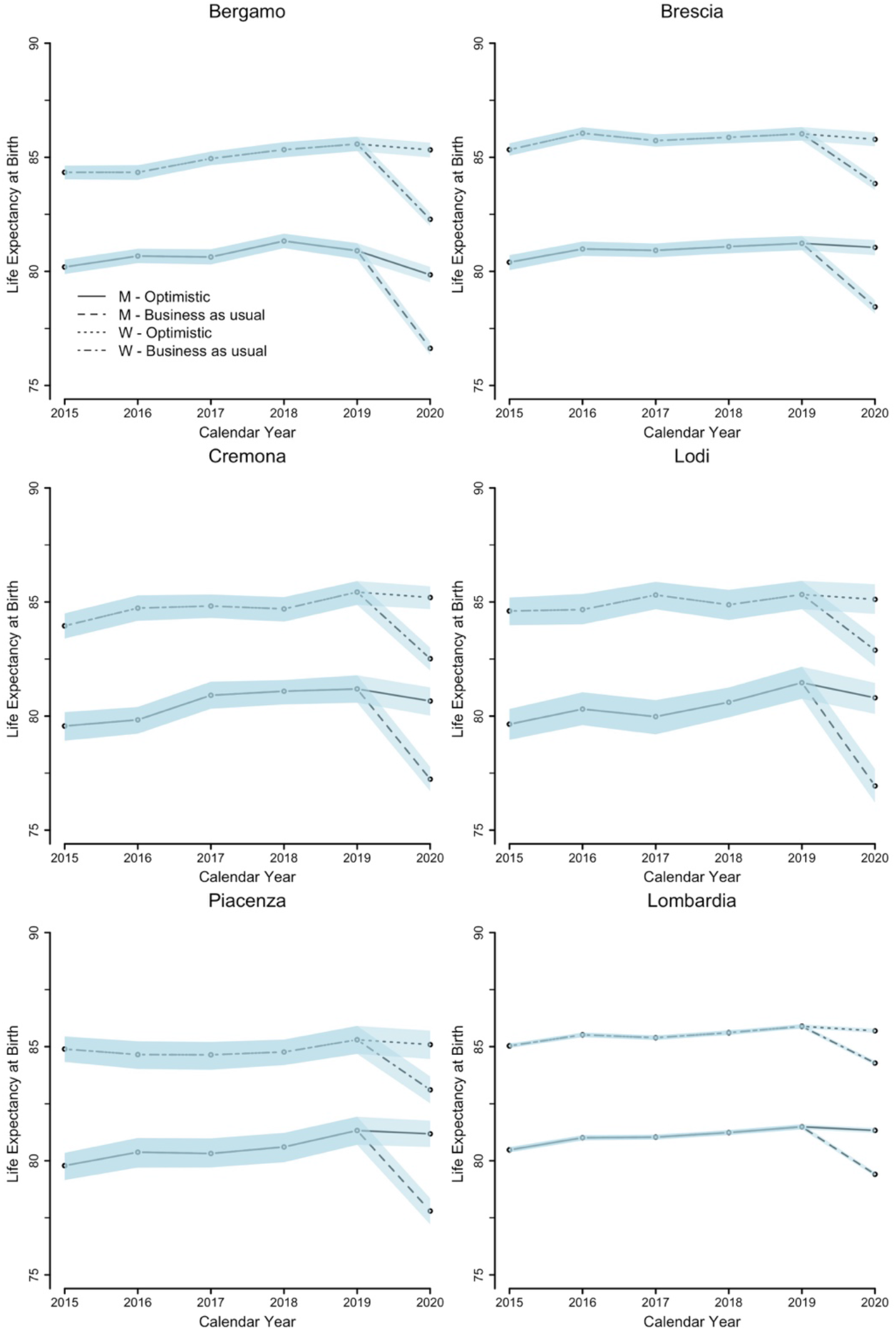
Estimates of annual life expectancy at birth by sex in selected provinces and in Lombardy (95% confidence intervals). Two scenarios: (A) optimistic scenario; and (B) business as usual scenario. Confidence intervals (95%) for life expectancies are estimated by bootstrapping using Monte Carlo simulation methods, assuming death counts follow a binomial distribution

When considering the first quadrimester of 2020, it is evident that the drop in life expectancy was significant for both men and women in all provinces (Figure 3). When compared to the average life expectancy of the 2015-2019 period, the years of life lost for men range from 6.2 years in Piacenza to 8 years in Bergamo and for women from 3.6 years in Piacenza to the 5.8 years in Bergamo. The higher years of life lost for men is due to sex differentials in COVID-19 mortality risk, as consistently found in both the official case fatality data ^39,40^ and our death registration data.

Indeed, when decomposing the loss in life expectancy to identify which age groups mainly contribute to a reduction in life expectancy (Figure S4 in SI Appendix), it is clear that the older populations, especially men aged 60-79 years play a major role.

When life expectancy is extrapolated for the whole year (Figure 4), the loss in life expectancy is naturally diluted over a longer period. The drop in life expectancy due to COVID-19-related excess mortality thus is less steep than that observed in the first quadrimester life expectancy (Figure 3). In the optimistic scenario of full harvesting (Figure 4), not much changes in life expectancy are expected with respect to the previous years. This rather unlikely situation represents a potential upper bound of life expectancy. Note however that in some cases, annual life expectancy in 2020 drops even in this optimistic scenario (for example, for men in Bergamo). This is because some age groups had already experienced in the first four months of 2020 more deaths than in the entire 2019 (see Figures S3A and S3B in SI Appendix). For these groups, mortality for 2020 is thus likely to be higher than what experienced in the previous years, even if full harvesting is assumed.

If mortality goes back to the pattern of normal times (‘business-as-usual’ scenario) (Figure 4), in the most severely hit province of Bergamo, life expectancy will drop by 4.1 and 2.6 years for men and women respectively when compared to the years 2015-2019. In slightly less affected provinces of Brescia, Cremona, Lodi and Piacenza, the years of life lost for men are between 2.5 in Brescia and 3.5 in Lodi and for women between 1.7 in Piacenza to 2.2 in Cremona. As expected, Lombardy shows a smaller reduction in life expectancy of 1.6. and 1.2 years for males and females, respectively.

## Discussion

This study estimates excess mortality directly and indirectly linked to COVID-19. Avoiding the inconsistencies in classification of cause of deaths and in testing practices and by focusing on the five most severely affected areas in Italy where the first wave of the pandemic appeared to be over, the present analysis provides an assessment of the full impact of the first wave of COVID-19 on population health and human life.

Two empirical regularities clearly emerge regarding demographic differentials. First, the age gradient in excess mortality is steep and age represents the most evident risk factor for COVID-19 mortality ^33,41^. In Lombardy, men and women aged over 70 are 19 times and 47 times more likely to die than their counterparts aged under 70 and under 60, respectively. These patterns are replicated in all five provinces. Therefore, an area with a high proportion of older populations (e.g. 17% of the population was over 70 in Lombardy region in 2019) would suffer a higher burden of COVID-19 mortality ^42^. Second, the risk of dying for men within each province is consistently higher than for women for all age classes and provinces considered. Higher mortality rates consequently translate into a larger reduction in life expectancy for men than for women.

Although these data provide evidence of the severity of the first wave of the COVID 19 pandemic in Europe, a further measurement effort is needed, particularly for geographical and historical comparability purposes. In terms of life expectancy, we have shown two main sets of results. First, when focusing on 1 January to 30 April 2020, the reduction in the first quadrimester life expectancy, compared with the average of the years 2015-2019, was as high as 8 years for men and 5.8 years for women in Bergamo, respectively. Second, when the analysis is extended to the whole year, under the realistic assumption (business-as-usual scenario) that mortality rates from May onward will be back to ‘normal’, life expectancy is reduced by up to 4 years (for men in Bergamo). In fact, there remain significant uncertainties regarding the longer-term effects of the pandemic on health conditions; for instance, regarding patients recovering from COVID-19 with important co-morbidities and mental health issues ^43,44^ or pregnant women ^45,46^. Indirect physical and mental health consequences following changing socioeconomic conditions might also affect mortality patterns in 2020 ^47^. Therefore, if anything, our estimates of annual life expectancy for 2020 in the business-as-usual scenario with no harvesting might actually be on an optimistic side.

A careful attention should be paid for the interpretation of such figures. Formally, period life expectancy at birth represents the average life span of an individual living under the current mortality regime i.e. the mean age at death for a newborn who is subjected to today’s age-specific mortality rates over his/her full life course. However, mortality is rarely constant over time and therefore, period life expectancy is usually a poor indicator for the life span of an actual group of individuals ^48^. This holds true particularly for sudden shocks, such as epidemics, wars or natural disasters, which temporarily increase mortality. For this reason, life expectancy is a powerful tool for measuring the lost of life years (human cost) associated to an event. Yet, the presented figures should not be understood as estimates for the life length of any Italian living in the analysed provinces. Specifically, one could think about the COVID-related loss of life expectancy as the differences between the expected life length of two hypothetical newborns growing up in two different alternative worlds: one with regular pre-COVID mortality rates and one with COVID mortality rates as measured by the present analysis.

The difference between the quadrimester and the annual life expectancy estimations is based on the assumption that the five provinces will not witness a second epidemic wave in autumn 2020. A second wave would further consistently reduce the annual estimation of life expectancy, but would not affect the estimation of life lost specific for the first four months of the year. In this sense, the figures for the first four months can be considered as an estimation of the human life lost *for the first wave* of the COVID-19 epidemics in the affected provinces and will not be influenced by the evolution of mortality after April 2020.

The COVID-19 pandemic is often compared with the 1918 influenza pandemic (commonly known as ‘Spanish flu’) – the most severe pandemic in the last century. The COVID-19 pandemic brought about a large-scale loss in life expectancy from pandemics, second only to the 1918 Spanish flu, where life expectancy (for Italy) dropped by more than 15 years (See Figure S5 in SI Appendix) and for the USA, by 11.8 years as estimated by Noymer and Garenne ^49^. Fortunately, the direct impact of COVID-19 on human life seems to be smaller than the 1918 Spanish flu for many reasons. First of all, since the Spanish flu typically killed younger people, it reduces life expectancy much more than the COVID-19 which is particularly fatal for older populations. Second, the outbreak of the Spanish flu occurred at the same time as the First World War when no public health and preventive measures were adequately implemented. The coexistence of war and pandemic made the Spanish flu exceptionally lethal. Finally, during the period of the 1918 influenza pandemic, the medical knowledge and technology were much less advanced, with penicillin and other crucial pharmaceutical products not yet being available. It thus can be said that the years of life lost observed in the most severely affected areas of Italy, although not at the level achieved after the Spanish flu, are of a substantial scale given a much more advanced medical technology and the absence of war in the present day.

The timing and the explicit focus on a local context can be considered as the strength of this analysis. Given that the COVID-19 outbreaks are geographically concentrated, looking at a country level life expectancy is misleading and underestimates the actual impact of the pandemic. The present analysis represents the first attempt to provide an evidence-based assessment of the human life impact from a frontline COVID-19 outbreak. Measuring the human costs associated to COVID-19 is of extreme importance. It is a warning for other countries and for the future, and, more importantly, it informs about the risks that maybe underestimated in favour of the economic aspects. While economic loss associated to the COVID-19 epidemic is often quantified in terms of GDP loss, here we offer the first quantification of human cost associated with COVID-19. Indeed, this type of information is highly sought in the public debate.

Human life in itself is a solid indicator of human development because it represents health and wellbeing of a population ^19,20^, which is one key priority of the Sustainable Development Goals. So many years of life lost due to COVID-19 pandemic is worrisome since it sets the affected areas back to their level of human development 15 years ago. As shown in the cases of Bergamo, Brescia, Cremona, Lodi, and Piacenza, the cost of human life from a delay in public interventions to reduce the virus transmission is disturbingly high. Amidst countries’ preparation to ease the lockdown restrictions and social distancing measures, it is important to keep in mind the conceivable risk of viral reintroduction and a potential direct and indirect cost it can pose on human life. Well-planned government measures to prevent a second wave of infections along with public collaboration in keeping physical distancing and practicing proper hygiene until a vaccine for the novel coronavirus is available are key to achieve a balance between health and economic protection.

## Data Availability

All data referred in the manuscript are freely accessible.
Data from the Italian National Institute of Statistics are made available on the Istat website: https://www.istat.it/
Data from the Human Mortality Database can be accessed at https://www.mortality.org/

## Acknowledgments

We would like to thank the Italian National Statistical Institutes (ISTAT) for providing invaluable assistance with data access and enquiries.

